# Vaccine Effectiveness Against Hospitalization Among Adolescent and Pediatric SARS-CoV-2 Cases in Ontario, Canada

**DOI:** 10.1101/2022.03.24.22272919

**Authors:** Alison E. Simmons, Afia Amoako, Alicia A. Grima, Kiera R. Murison, Sarah A. Buchan, David N. Fisman, Ashleigh R. Tuite

**Affiliations:** Dalla Lana School of Public Health, University of Toronto, Toronto, Ontario, Canada; Public Health Ontario, Toronto, Ontario, Canada; Centre for Immunization Readiness, Public Health Agency of Canada, Ottawa, Ontario, Canada

**Keywords:** SARS-CoV-2, vaccination, hospitalization, pediatric, adolescent

## Abstract

**Background:** Vaccines against SARS-CoV-2 have been shown to reduce risk of infection, as well as severe disease among those with breakthrough infection, in adults. The latter effect is particularly important as Immune evasion by Omicron variants appears to have made vaccines less effective for prevention of infection. There is currently little available information on the protection conferred by vaccination against severe illness due to SARS-CoV-2 in children.

**Methods:** To minimize confounding by changing vaccination practices and dominant circulating viral variants, we performed an age- and time-matched nested case-control design. Reported SARS-CoV-2 case records in Ontario children and adolescents aged 4 to 17 were linked to vaccination records. We used multivariable logistic regression to estimate the effectiveness of one and two vaccine doses against hospitalization.

**Results:** We identified 130 hospitalized SARS-CoV-2 cases and 1,300 non-hospitalized, age- and time-matched controls, with disease onset between May 28, 2021 and January 9, 2022. One vaccine dose was shown to be 34% effective against hospitalization among SARS-CoV-2 cases (aOR = 0.66 [95% CI: 0.34, 1.21]). In contrast, two doses were 56% (aOR = 0.44 [95% CI: 0.23, 0.83]) effective at preventing hospitalization among SARS-CoV-2 cases. Exploratory instrumental variable analyses, and calculation of E-values, suggested that these effects are unlikely to be explained by unmeasured confounding.

**Conclusions:** Even with immune evasion by SARS-CoV-2 variants, two vaccine doses continue to provide protection against hospitalization among adolescent and pediatric SARS-CoV-2 cases, even when the vaccines do not prevent infection.

## Introduction

Severe Acute Respiratory Syndrome (SARS-CoV-2) has caused more than 5.5 million deaths globally (1). Safe and effective vaccines to prevent SARS-CoV-2 infection and severe outcomes have been approved since late 2020 for adults (2). With the emergence of B.1.617 (Delta) in May 2021 and B.1.1.529 (Omicron) in November 2021, decreased vaccine effectiveness against infection was observed (3-6). Given that the goal of vaccination is to prevent death, severe disease, and overall disease burden, it is important to consider how well vaccines achieve these goals among individuals infected with SARS-CoV-2 (7).

Four recent studies focused on quantifying the real-world effectiveness of SARS-CoV-2 vaccination against hospitalization among adolescent or pediatric populations (8-11). All the studies used uninfected controls, and resulting vaccine effectiveness estimates reflect the joint risk of SARS-CoV-2 infection and the risk of hospitalization conditional on infection (12). Olson et al.(8) showed that BNT162b2 was 94% effective against hospitalization among adolescents infected with the Delta variant (B.1.617). After the emergence of the Omicron variant (B.1.1.529), two dose vaccine effectiveness against hospitalization was estimated to be 73% among adolescents ages 12 to 17 and 48% among pediatric patients ages 5 to 11 (9). In a recent Morbidity and Mortality Weekly Report (MMWR), two dose vaccine effectiveness against hospitalization was between 73% and 94% among adolescent and pediatric populations (10). In a study among hospitalized patients, two dose vaccine effectiveness against SARS-CoV-2 hospitalization was 93% during the delta-predominant period and 40% during the omicron-dominant period among adolescents, and 68% during the omicron-dominant period among pediatric patients (11). In contrast to the approach used in these studies, we conditioned upon SARS-CoV-2 infection in our analysis to estimate the direct effect of vaccination on hospitalization risk among pediatric and adolescent SARS-CoV-2 cases (13).

The Canadian province of Ontario is a large (population 14.6 million) and diverse jurisdiction, with high levels of SARS-CoV-2 vaccine coverage (14). Approximately 87% of Ontario residents ages 12 to 17 and 53% of Ontario residents ages 5 to 11 received at least one SARS-CoV-2 dose as of January 2022 (15). Most individuals ages 12-17 were eligible to receive the BNT162b2 (Pfizer-BioNTech, “Comirnaty”) vaccine beginning on May 28, 2021, followed by ages 5 to 11 on November 28, 2021 (16). Robust public health surveillance systems in Ontario enable individual-level linkage of the SARS-CoV-2 vaccination database and the reportable disease database. Data include individual-level demographic factors that can be used to reduce bias in our vaccine effectiveness estimates. We calculated one dose and two dose vaccine effectiveness against hospitalization among adolescent and pediatric SARS-CoV-2 cases

## Methods

### Data Sources

Confirmed SARS-CoV-2 cases were identified in Ontario’s Public Health Case and Contact Management Solution (CCM). The CCM includes patient demographics (i.e., sex, age, comorbidities), geographic location (i.e., public health unit), and case characteristics (i.e., test date, symptom onset date, hospital admission and discharge dates) for all laboratory-confirmed SARS-CoV-2 cases in Ontario. SARS-CoV-2 vaccination information was identified from the provincial COVaxON database. COVaxON data includes vaccine administration information (i.e., dose dates, dose locations, dose indication, vaccine product) for Ontario residents with a provincial health insurance number. The CCM and COVaxON data were linked through a unique “pseudo-health card number” identifier present in both datasets. Median household income at the census subdivision level was from the 2016 Census of Population (17).

### Study Design

We conducted a nested case-control study from a cohort of individuals aged 4-17 years who had a confirmed SARS-CoV-2 infection in Ontario, Canada. These individuals had a positive reverse transcription real-time polymerase chain reaction (PCR) test between May 28, 2021 and January 9, 2022. Each hospitalized SARS-CoV-2 case was matched to 10 non-hospitalized SARS-CoV-2 cases by case onset date and age to improve precision (18). Case onset date was defined as the date of symptom onset among symptomatic cases and the specimen collection date for SARS-CoV-2 testing among those without a symptom onset date.

We restricted our analysis to incident SARS-CoV-2 infections (re-infections were excluded). We excluded SARS-CoV-2 cases where individuals had received a vaccine not approved for use in this population in Canada (e.g., Johnson & Johnson/Janssen COVID-19 vaccine, Oxford-AstraZeneca COVID-19 vaccine) or had received three vaccine doses, as this population was not eligible for third doses (i.e., booster dose) during the study period. We extracted the data on January 19, 2022, but only included test dates up to January 9, 2022, to account for delays between testing, hospitalization, and reporting. SARS-CoV-2 cases aged 4 years were included in the analysis because cases among pediatric patients aged 4 and aged 5 were in the same 2-year age group in these data, and pediatric patients who turned 5 in the 2021 calendar year were eligible for vaccination. In total, all 130 hospitalized SARS-CoV-2 cases were matched with 1,300 non-hospitalized SARS-CoV-2 cases (**Figure 1**).

**Figure 1.**
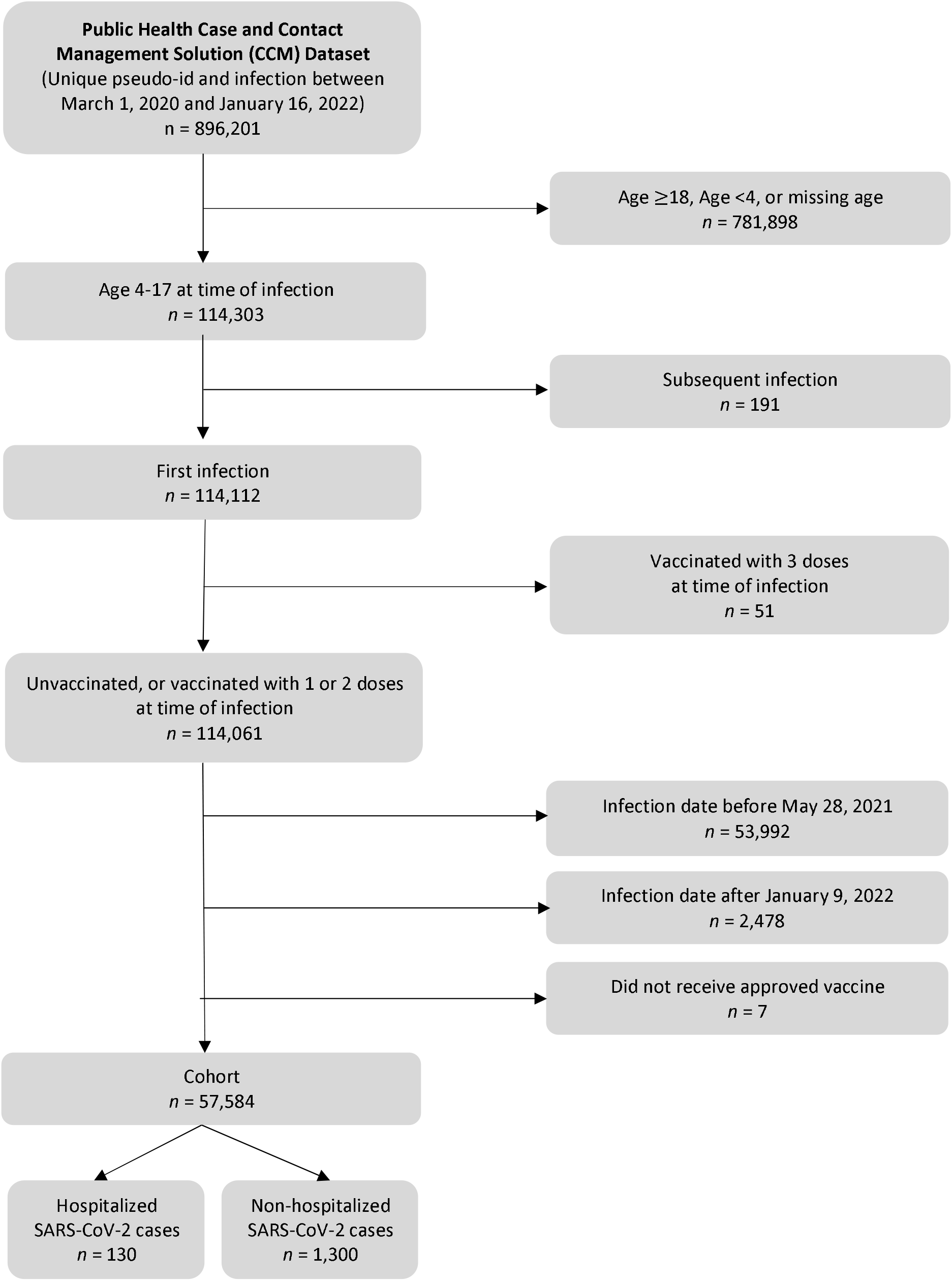
Flow Diagram for Creation of Nested Case-Control Study Population

### Measures

The outcome was hospitalization due to SARS-CoV-2. Hospitalizations were identified by a reported hospital admission date, or a reported hospitalization or ICU admission due to SARS-CoV-2 in the CCM. The exposure was SARS-CoV-2 vaccination. We considered individuals one dose vaccinated 14 or more days after the date the first vaccine dose was administered; individuals were considered two dose vaccinated 14 or more days after the date the second vaccine dose was administered. During the first 13 days after the date the first vaccine dose was administered, individuals were considered unvaccinated.

Case onset date, age, sex, asthma, and immunocompromising condition were included as confounders (11, 19). A Directed Acyclic Graph (DAG) outlining the main hypothesized relationships between the variables is presented in **Figure S1** (20). The proportion of two dose vaccinated individuals less than age 18 years within each public health unit was included as an instrumental variable in a sensitivity analysis. Additionally, median household income at the census subdivision level was included as a confounder in a sensitivity analysis.

### Statistical Analysis

We calculated frequencies of baseline characteristics and compared SARS-CoV-2 cases by hospitalization status. Standardized mean differences (SMD) were used in addition to statistical tests (Chi-square and unpaired t-tests) to compare differences in covariates between hospitalized and non-hospitalized groups. Standardized mean differences are not impacted by sample size, in contrast to the common statistical tests used in descriptive tables. A standardized difference greater than 0.10 represents a lack of covariate balance (21).

We used multivariable logistic regression to calculate the effectiveness of one and two vaccine doses against hospitalization among adolescent (ages 12-17) and pediatric (ages 4-11) SARS-CoV-2 cases, while adjusting for sex, asthma, immunocompromising condition, age, and case onset date. Subsequently, we examined potential effect measure modification by age group and case onset date. To assess effect measure modification by age-group, we used two multivariable logistic regression models to separately calculate vaccine effectiveness among those ages 4-11 and ages 12-17 while adjusting for sex, asthma, immunocompromising condition, age, and case onset date. To assess effect measure modification by infecting variant (i.e., primarily Delta and Omicron), we included an interaction term between vaccination status and case onset data (May 28, 2021 – December 14, 2021 compared to December 15, 2021 – January 9, 2022) in a multivariable logistic regression model among adolescents. Ontario residents under age 12 were ineligible for SARS-CoV-2 vaccination during most of the former time-period and were thus excluded from the analysis. Dates were chosen to align with the rise in the prevalence of the Omicron variant in Ontario. Vaccine effectiveness (VE) was calculated using the formula VE = (1-aOR)*100%.

We performed sensitivity analyses to quantify and mitigate the impact of unmeasured confounding. First, we calculated an E-value based on the results of the non-instrumented multivariable logistic regression model (22). An E-value quantifies how strong an unmeasured confounder would need to be to explain away the association between SARS-CoV-2 vaccination and hospitalization (23). Second, we used instrumented-multivariable logistic regression to estimate the average impact of two doses of vaccination on SARS-CoV-2 hospitalization risk with additional control for unmeasured confounders (24). We used the proportion of individuals in each age group (adolescent or pediatric) vaccinated with two doses of SARS-CoV-2 vaccine in each public health unit (hereby referred to as PHU vaccine proportion) on January 9, 2022 as a measure of vaccine acceptance, and as an instrument to predict individual-level vaccination.

Instrumental variable methods can be used to estimate the average causal effect of vaccination on hospitalization in the presence of unmeasured confounders under the following assumptions: (i) the instrument causes the exposure, (ii) affects the outcome only through the exposure, and (iii) shares no common causes with the outcome (24). We verified the first assumption by quantifying the association between PHU vaccine proportion and vaccination status (*r*^2^=0.31), and the latter two assumptions based on theory and our DAG (**Figure S1**). We used a two-stage model to calculate the adjusted odds ratio (aOR) of the average impact of 2-dose SARS-CoV-2 vaccination on hospitalization under the assumption of homogeneity (i.e., no effect measure modification) (25). The first model used multivariable probit regression to calculate predicted levels of vaccination conditional on PHU vaccine proportion and the measured confounders. The second model used logistic regression with hospitalization as the outcome, and the predicted levels of vaccination from the probit model and the measured confounders as predictors. Robust standard errors were calculated with the Sandwich estimator (26).

We also performed additional sensitivity analyses to explore the robustness of our results to our assumptions. First, we included median household income at the census subdivision level as a confounder in addition to sex, asthma, immunocompromising condition, age, and case onset date in a multivariable logistic regression model. Individual-level measures of socioeconomic status were not available in our data and is therefore an unmeasured confounder in our study (11). Second, we excluded SARS-CoV-2 cases with case onset dates within 13 days of the administration of the first or second vaccine dose. In our main analysis, we classified SARS-CoV-2 cases as unvaccinated for the first 13 days after dose one was administered and as one dose vaccinated for the first 13 days after dose two was administered. Last, we excluded SARS-CoV-2 cases with case onset dates after January 5, 2022 to allow for a 14-day delay between case onset date and hospitalization. In our main analysis, we allowed for a 10-day delay between case onset date and hospitalization.

### Ethics

We received ethics approval for this study from the Research Ethics Board at the University of Toronto (#00031358).

## Results

We included 130 hospitalized SARS-CoV-2 cases and 1,300 nonhospitalized SARS-CoV-2 cases in our analysis, with test report dates between May 28, 2021 and January 9, 2022 (**Figure 1**). Individuals hospitalized with SARS-CoV-2 were more likely to be unvaccinated, immunocompromised, and have an asthma diagnosis compared to those who were not hospitalized. The distribution of age and case onset date did not differ by hospitalization status due to the age and time-matched study design (**Table 1**).

**Table 1.** Description of adolescent and pediatric SARS-CoV-2 cases by hospitalization status (*n* = 1,430)

| Characteristic | Hospitalized<br>$n = 130$ | | Non-hospitalized<br>$n = 1,300$ | | SMD | $p^a$ |
| --- | --- | --- | --- | --- | --- | --- |
| | $n$ | (%) | $n$ | (%) | | |
| Vaccination Status<sup>b</sup> |  |  |  |  | 0.19 | 0.18 |
| Unvaccinated | 99 | (76.2) | 893 | (68.7) |  |  |
| One dose | 14 | (10.8) | 164 | (12.6) |  |  |
| Two doses | 17 | (13.1) | 243 | (18.7) |  |  |
| Male |  |  |  |  | 0.05 | 0.67 |
| Yes | 66 | (50.8) | 691 | (53.2) |  |  |
| No | 64 | (49.2) | 609 | (46.8) |  |  |
| Immunocompromised |  |  |  |  | 0.43 | <0.001 |
| Yes | 12 | (9.2) | <5 | (<0.4) |  |  |
| No | 118 | (90.8) | >1,295 | (>99.6) |  |  |
| Asthma |  |  |  |  | 0.27 | <0.001 |
| Yes | 8 | (6.2) | 15 | (1.2) |  |  |
| No | 122 | (93.8) | 1,285 | (98.8) |  |  |
|  | **Mean** | **(SD)** | **Mean** | **(SD)** |  |  |
| Age | 11.2 | (4.3) | 11.0 | (4.3) | 0.05 | 0.59 |
| Case Onset Date<sup>c</sup> | 2021-10-17 | (79.0 days) | 2021-10-16 | (79.7 days) | 0.01 | 0.91 |

Notes: SMD = standardized mean difference; $p$ = p-value; SD = standard deviation; Vaccination status is the exposure; Male, immunocompromised status, and asthma are the covariates; Age and case onset date are the matching factors
<sup>a</sup> Pearson chi-square for categorical variables, and unpaired t-tests for continuous variables
<sup>b</sup> Vaccination status on case onset date
<sup>c</sup> Date of symptom onset for symptomatic cases and the specimen collection date for asymptomatic cases

In our main analysis, after adjustment for sex, asthma, immunocompromising condition, age, and case onset date, we observed that one SARS-CoV-2 vaccine dose was 34% effective (aOR = 0.66 [95% CI: 0.34, 1.21]) against hospitalization among adolescent and pediatric SARS-CoV-2 cases (**Table 2**). In contrast, two SARS-CoV-2 vaccine doses were 56% effective (aOR = 0.44 [95% CI: 0.23, 0.83]) at preventing hospitalization among SARS-CoV-2 cases. In our stratified analysis, we found that one SARS-CoV-2 vaccine dose was 31% effective (adjusted odds ratio (aOR) = 0.69 [95% CI: 0.22, 1.78]) and two SARS-CoV-2 vaccine doses were 58% effective (adjusted odds ratio (aOR) = 0.42 [95% CI: 0.20, 0.84]) at preventing hospitalization among adolescent patients ages 12-17 years. One SARS-CoV-2 vaccine dose 34% effective (aOR = 0.66 [95% CI: 0.27, 1.45]) at preventing hospitalization among pediatric SARS-CoV-2 cases ages 4-11. We did not find statistically significant effect measure modification by case onset date among SARS-CoV-2 cases ages 12-17 (likelihood ratio test, *p*=0.34) (**Figure S2**).

**Table 2.** Adjusted odds ratios, with 95% confidence intervals, of the relationship between vaccination status and hospitalization among adolescent and pediatric SARS-CoV-2 cases (*n* = 1,430)

|  | **Overall**<br>*n* = 1,430 |  | **Ages 12-17**<br>*n* = 673 |  | **Ages 4-11**<br>*n* = 757 |  |
| --- | --- | --- | --- | --- | --- | --- |
| **Characteristic** | **aOR** | **(95% CI)** | **aOR** | **(95% CI)** | **aOR** | **(95% CI)** |
| Vaccination Status<sup>a</sup> |  |  |  |  |  |  |
| Unvaccinated | 1.00 | (ref) | 1.00 | (ref) | 1.00 | (ref) |
| One dose | 0.66 | (0.34, 1.21) | 0.69 | (0.22, 1.78) | 0.66 | (0.27, 1.45) |
| Two doses | 0.44 | (0.23, 0.83) | 0.42 | (0.20, 0.84) | --<sup>b</sup> | -- |
| Male |  |  |  |  |  |  |
| Yes | 0.86 | (0.59, 1.25) | 0.82 | (0.47, 1.41) | 0.91 | (0.53, 1.55) |
| No | 1.00 | (ref) | 1.00 | (ref) | 1.00 | (ref) |
| Immunocompromised |  |  |  |  |  |  |
| Yes | 50.80 | (15.64, 277.57) | 73.30 | (11.80, 142.04) | 38.2 | (8.36, 269.46) |
| No | 1.00 | (ref) | 1.00 | (ref) | 1.00 | (ref) |
| Asthma |  |  |  |  |  |  |
| Yes | 6.31 | (2.46, 15.06) | 5.95 | (1.51, 20.62) | 6.49 | (1.66, 21.95) |
| No | 1.00 | (ref) | 1.00 | (ref) | 1.00 | (ref) |

Notes: aOR = adjusted odds ratio; CI = confidence interval; ref = reference; Adjusted for sex, immunocompromised status, asthma, age, and case onset date
<sup>a</sup> Vaccination status on case onset date
<sup>b</sup> 5 SARS-CoV-2 cases occurred among pediatric patients ages 4-11 vaccinated with two doses, and <5 patients were hospitalized

In our primary analysis, the E-value for the adjusted association between vaccination with two doses and the risk of hospitalization (i.e., aOR = 0.44) was 4.31 (22). An unmeasured confounder would need to be independently associated with vaccination and hospitalization by a 4.31-fold risk ratio to result in a null association, after adjusting for sex, asthma, immunocompromising condition, age, and case onset date. Similarly, the E-value for the upper 95% CI bound (i.e., 95% CI upper bound = 0.83) is 1.70. A set of unmeasured confounders would need to be independently associated with vaccination and hospitalization by a 1.70-fold risk ratio for the 95% CI to encompass the null value after controlling sex, asthma, immunocompromising condition, age, and case onset date. In our instrumental variable analysis, which accounts for unmeasured confounding, two SARS-CoV-2 vaccine doses were 38% effective (adjusted OR (aOR) = 0.62 [95% CI: 0.22, 1.79]) at preventing hospitalization among adolescent SARS-CoV-2 cases (**Table S1**).

Our vaccine effectiveness estimates did not substantially change when included income at the census subdivision level, when we excluded SARS-CoV-2 cases with case onset dates within 13 days of receiving a first or second vaccine dose, and when case onset dates after January 5, 2022 were excluded (**Table S2; Table S3; Table S4**).

## Discussion

Among adolescent and pediatric SARS-CoV-2 cases, one vaccine dose was 34% effective, and two vaccine doses were 56% effective at preventing SARS-CoV-2 hospitalization. We found no evidence of effect modification by age-group. One SARS-CoV-2 vaccine dose was 31% effective, and two SARS-CoV-2 doses were 58% effective at preventing hospitalization among cases ages 12-17 years, and one SARS-CoV-2 vaccine dose was 34% effective at preventing hospitalization among cases ages 4-11 years. The effectiveness of two SARS-CoV-2 vaccine doses could not be examined independently in the pediatric population due to a small number of two-dose vaccinated hospitalized cases. We did not evidence of effect measure modification by case onset date. Our vaccine effectiveness estimates are robust to unmeasured confounding, as evidenced by the high E-value for the estimates. Similarly, two doses of SARS-CoV-2 vaccine remained effective against hospitalization after additional control for unmeasured confounding in our instrumental variable analysis. Our study demonstrates that among adolescent and pediatric SARS-CoV-2 cases, vaccination provides additional protection against hospitalization independent of protection provided against infection.

The studies that have focused on SARS-CoV-2 vaccine effectiveness against hospitalization in adolescent and pediatric populations are not directly comparable to ours due to differences in control selection. Our estimated two dose vaccine effectiveness estimates against hospitalization are lower than most estimates from studies using negative controls in the United States (8-10), likely because we isolated the impact of vaccination of hospitalization risk, independent of SARS-CoV-2 susceptibility. In a case-control study conducted between May and October 2021 (i.e., prior to widespread infection with Omicron) with test-negative and syndrome-negative controls, vaccine effectiveness against hospitalization was 94% among those with two doses and 97% among those with one dose (8). A study with 164 hospitals in the United States from April 2021 to January 2022 found that two dose vaccine effectiveness against hospitalization was 74% among children ages 5-11 years and between 73% and 94% among adolescents (10). In contrast, two dose vaccine effectiveness was calculated to be 40% against hospitalization among adolescents during the circulation of omicron in a study including 31 hospitals across the United States (11). This lower estimate but may be due to earlier adolescent vaccination dates in the United States compared to in Ontario, as well as the impact of vaccine waning (11). Unlike prior studies, our control group consisted of non-hospitalized SARS-CoV-2 cases, not uninfected individuals. This allowed for the isolation of the effectiveness of vaccination against hospitalization independent of infection risk, which explains our lower vaccine effectiveness estimates against hospitalization compared to the majority of prior studies (12). Related, we were able to match our non-hospitalized and hospitalized cases on case onset date to maximize our ability to control for time, and thus circulating SARS-CoV-2 variant (18).

Our study has several notable strengths. Due to Ontario’s high quality public health surveillance data, we isolated the direct impact of vaccination on hospitalization risk with control for individual level demographic and health related factors. Our study includes a diverse population in a region with publicly funded healthcare (27, 28). We used a quantitative bias analysis to demonstrate the susceptibility of our results to unmeasured confounding (22), and we conducted used instrumental variable methods to adjust for unmeasured confounders (24). Our study also had a few limitations. First, we did not consider time since vaccination in our analysis. In a recent study in the United States, time since vaccination was not shown to significantly impact SARS-CoV-2 vaccine effectiveness estimates against hospitalization in this population (10). We were unable to assess two dose effectiveness against hospitalization specifically among cases ages 4-11 years due to few hospitalizations among two-dose vaccinated individuals. The availability of testing, and the propensity to get tested may have differed between vaccinated and unvaccinated individuals, and the reporting of comorbidities may have been more common among hospitalized SARS-CoV-2 cases. We used time as proxy for infecting variant among adolescents in our evaluation of effect measure modification which may lead to misclassification. Finally, our number of hospitalized SARS-CoV-2 cases was small, and there may have insufficient power and precision to reliably assess effect measure modification by age and time.

With the continued emergence of variants that may further decrease SARS-CoV-2 vaccine effectiveness against infection, it is vital to consider how effective these vaccines are at preventing severe outcomes (29). Given that two dose vaccine effectiveness against hospitalization following infection is 58% in adolescents, and may be lower in youth ages 4-11 years, it is important to continue pharmaceutical and non-pharmaceutical interventions to prevent infection risk in these populations. Given that SARS-CoV-2 is primarily an airborne infection, this study supports the continued use of high-quality masks, increased ventilation, and accessible rapid antigen tests in addition to vaccination (30, 31).

## Conclusions

In this evaluation of the effectiveness of SARS-CoV-2 vaccination in adolescent and pediatric cases ages 4 to 17 in Ontario, Canada, we found that vaccination provides protection against hospitalization, even when the vaccines do not prevent infection. SARS-CoV-2 vaccines remain an effective intervention to prevent severe outcomes in adolescent and pediatric populations, and continued efforts are needed to increase vaccine uptake in these populations.

## Data Availability

Data used are property of the Government of Ontario and cannot be provided by the authors.

## Acknowledgements

We wish to thank the staff at Public Health Ontario and Ontario’s public health units for collecting, sequencing, analyzing, and providing access to the data used for this analysis.

## Supporting Information

**Figure S1.**
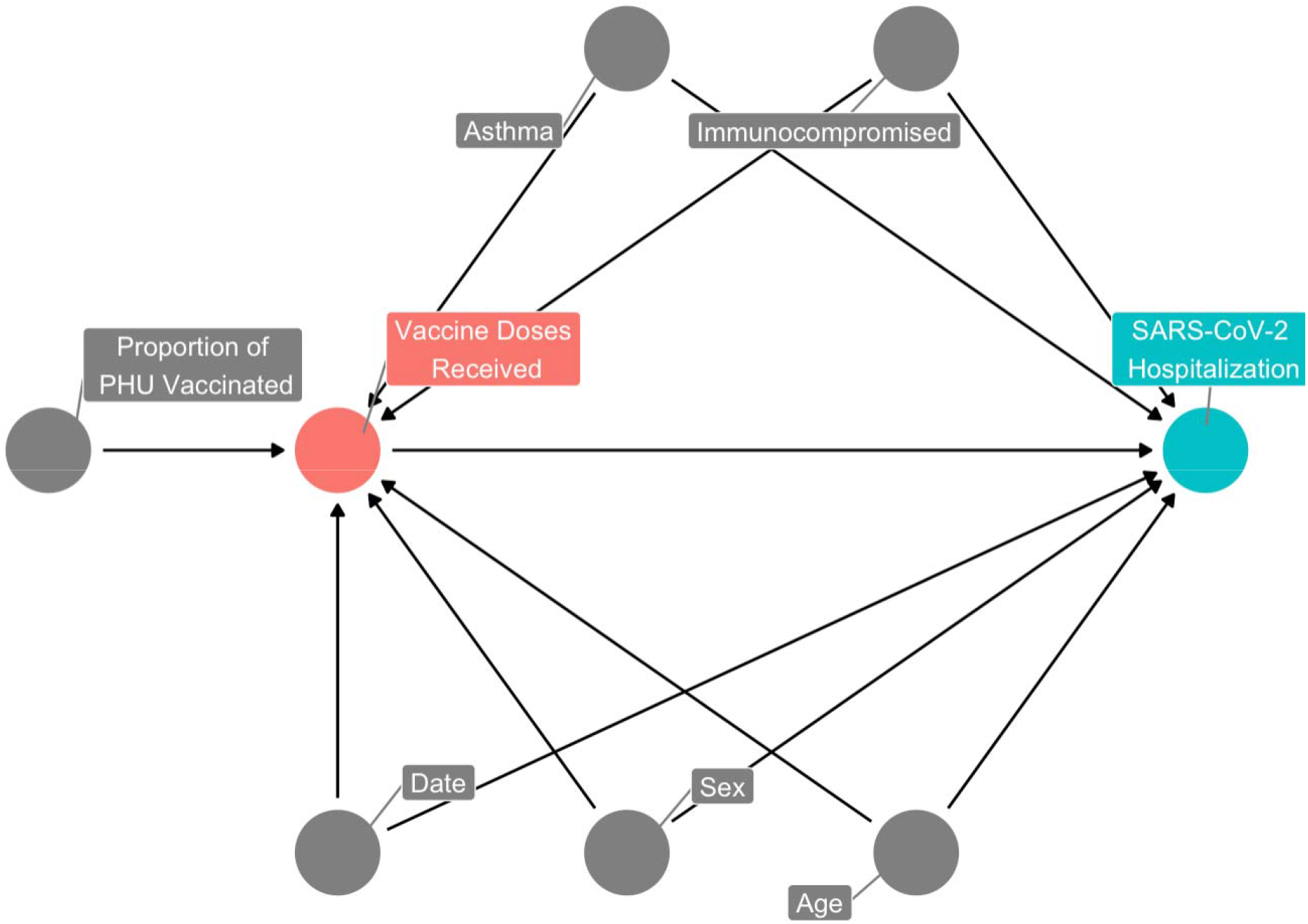
Directed Acyclic Graph (DAG) of the relationship between vaccination status and hospitalization among pediatric SARS-CoV-2 cases

**Figure S2.**
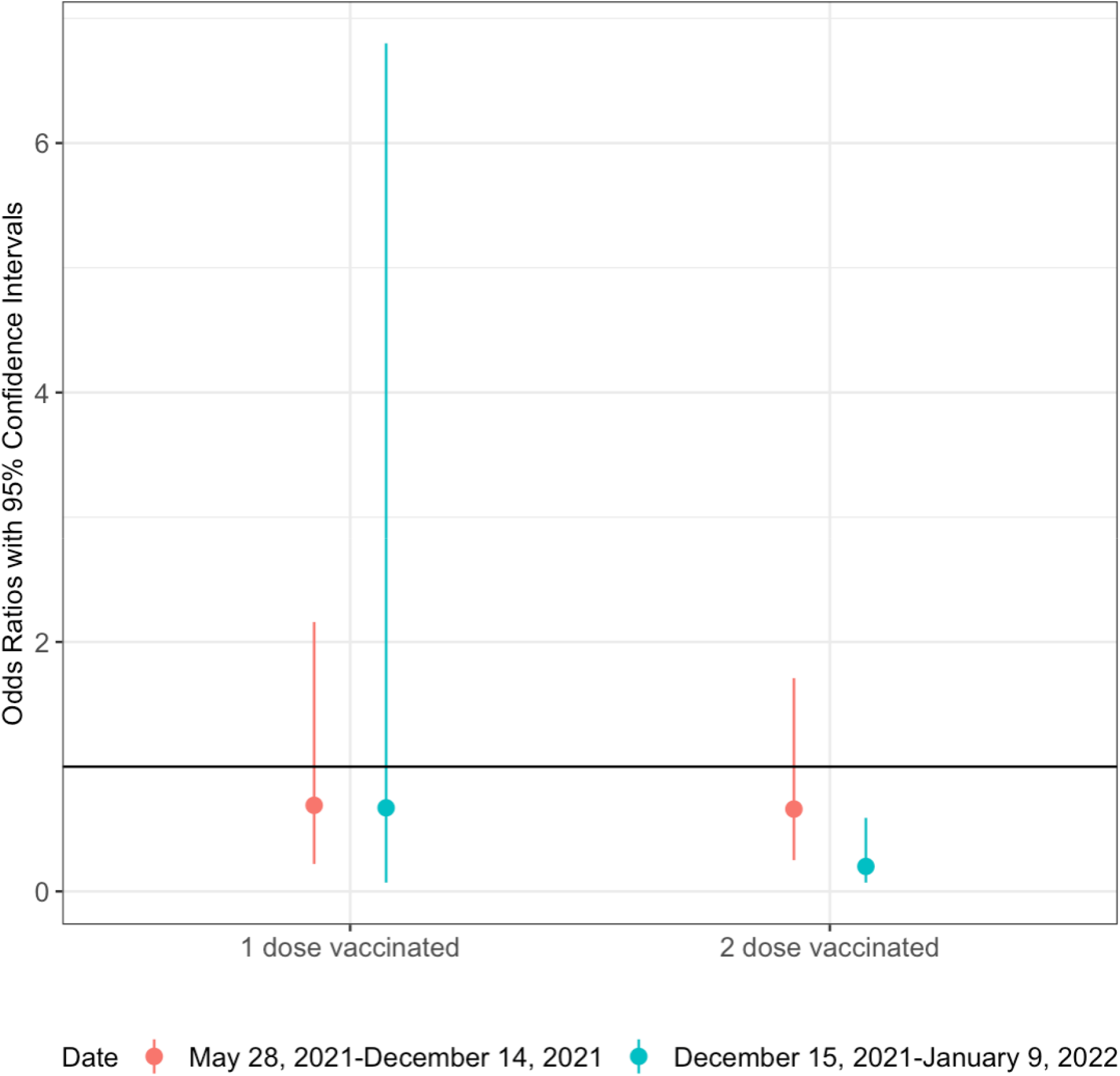
Adjusted odds ratios, with 95% confidence intervals, of the association between vaccination status and hospitalization among SARS-CoV-2 cases ages 12-17 stratified by vaccination status and case onset date (*n* = 673)

**Table S1.** Effect of vaccination on hospitalization among adolescent and pediatric SARS-CoV-2 cases using instrumental-variable adjusted regression with robust standard errors (*n* = 1,252)

| Characteristic | aOR | (95% CI) |
| --- | --- | --- |
| Vaccination Status<sup>a</sup> |  |  |
| Unvaccinated | 1.00 | (ref) |
| Two doses | 0.62 | (0.22, 1.79) |
| Male |  |  |
| Yes | 0.81 | (0.54, 1.20) |
| No | 1.00 | (ref) |
| Immunocompromised |  |  |
| Yes | 41.45 | (10.60, 162.04) |
| No | 1.00 | (ref) |
| Asthma |  |  |
| Yes | 6.80 | (2.83, 16.37) |
| No | 1.00 | (ref) |

*Notes:* aOR = adjusted odds ratio; CI = confidence interval; ref = reference; Adjusted for sex, immunocompromised status, asthma, age, and case onset date
<sup>a</sup> Vaccination status on case onset date

**Table S2.** Adjusted odds ratios, with 95% confidence intervals, of the relationship between vaccination status and hospitalization among adolescent and pediatric SARS-CoV-2 cases with census subdivision level income (1,393)

| Characteristic | aOR | (95% CI) |
| --- | --- | --- |
| Vaccination Status<sup>a</sup> |  |  |
| Unvaccinated | 1.00 | (ref) |
| One dose | 0.74 | (0.30, 1.32) |
| Two doses | 0.48 | (0.24, 0.90) |
| Male |  |  |
| Yes | 0.83 | (0.58, 1.26) |
| No | 1.00 | (ref) |
| Immunocompromised |  |  |
| Yes | 44.7 | (15.70, 228.64) |
| No | 1.00 | (ref) |
| Asthma |  |  |
| Yes | 6.66 | (2.88, 21.91) |
| No | 1.00 | (ref) |
| Income<sup>b</sup> | 1.00 | (1.00, 1.00) |

Notes: aOR = adjusted odds ratio; CI = confidence interval; ref = reference; Adjusted for sex, immunocompromised status, asthma, area-level income, age, and case onset date
<sup>a</sup> Vaccination status on case onset date
<sup>b</sup> Median household income at the census subdivision level

**Table S3.** Adjusted odds ratios, with 95% confidence intervals, of the relationship between vaccination status and hospitalization among adolescent and pediatric SARS-CoV-2 cases. SARS-CoV-2 cases with case onset dates within 13 days of their dose 1 or dose 2 administration date were excluded (*n* = 1,364)

| Characteristic | aOR | (95% CI) |
| --- | --- | --- |
| Vaccination Status<sup>a</sup> |  |  |
| Unvaccinated | 1.00 | (ref) |
| One dose | 0.66 | (0.30, 1.32) |
| Two doses | 0.47 | (0.24, 0.90) |
| Male |  |  |
| Yes | 0.86 | (0.58, 1.26) |
| No | 1.00 | (ref) |
| Immunocompromised |  |  |
| Yes | 51.01 | (15.70, 228.64) |
| No | 1.00 | (ref) |
| Asthma |  |  |
| Yes | 8.16 | (2.88, 21.91) |
| No | 1.00 | (ref) |

Notes: aOR = adjusted odds ratio; CI = confidence interval; ref = reference; Adjusted for sex, immunocompromised status, asthma, age, and case onset date
<sup>a</sup> Vaccination status on case onset date

**Table S4.**
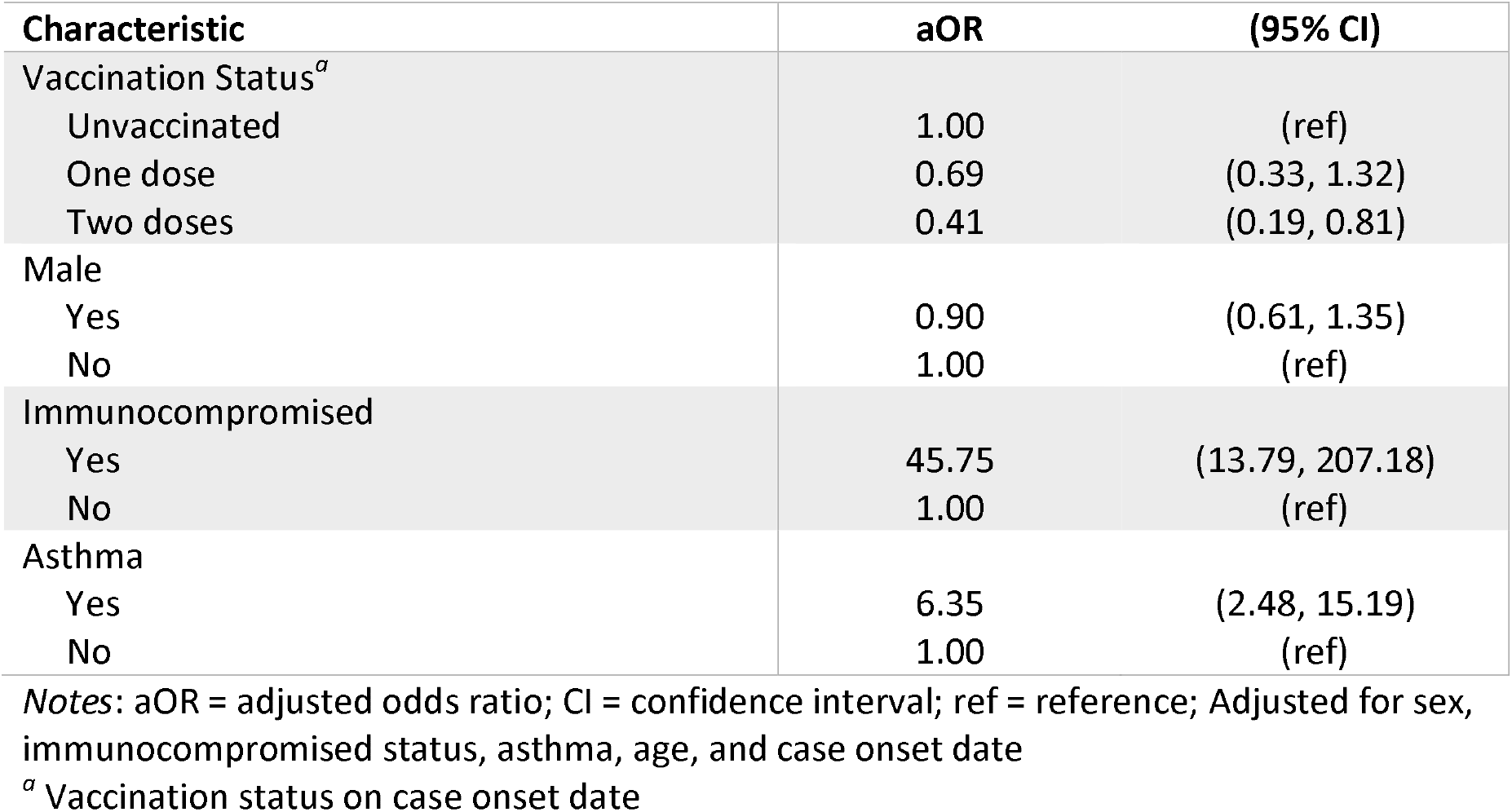
Adjusted odds ratios, with 95% confidence intervals, of the relationship between vaccination status and hospitalization among adolescent and pediatric SARS-CoV-2 cases. SARS-CoV-2 cases with case onset dates after January 5, 2022 were excluded (*n* = 1,364)

## Notes

**Funding** This work is supported by the University of Toronto [Connaught International Doctoral Scholarship to AES] and the Canadian Institutes of Health Research [2019 COVID-19 rapid research funding OV4-170360 to DNF].

### Competing Interest Statement

DNF has served on advisory boards related to influenza and SARS-CoV-2 vaccines for Seqirus, Pfizer, Astrazeneca and Sanofi-Pasteur Vaccines, and has served as a legal expert on issues related to COVID-19 epidemiology for the Elementary Teachers Federation of Ontario and the Registered Nurses Association of Ontario.
ART was employed by the Public Health Agency of Canada when the research was conducted. The work does not represent the views of the Public Health Agency of Canada.

### Funding Statement

AES is supported by the University of Toronto Connaught International Doctoral Scholarship. This research is supported by a grant to DNF from the Canadian Institutes for Health Research (2019 COVID-19 rapid researching funding OV4-170360).

### Summary of Updates

Updated dataset contains 130 rather than 131 cases; 10 fewer controls. Results change slightly. Clearer description of robustness and IV analysis results.

